# Protocol of a population-based prospective COVID-19 cohort study Munich, Germany (KoCo19)

**DOI:** 10.1101/2020.04.28.20082743

**Authors:** Katja Radon, Elmar Saathoff, Michael Pritsch, Jessica Michelle Guggenbühl Noller, Inge Kroidl, Laura Olbrich, Verena Thiel, Max Diefenbach, Friedrich Riess, Felix Forster, Fabian Theis, Andreas Wieser, Michael Hoelscher, the KoCo19 collaboration group^#^

**Affiliations:** Institute and Outpatient Clinic for Occupational, Social and Environmental Medicine, Ludwig Maximilian University (LMU) University Hospital Munich, Ziemssenstr. 1, D-80336 Munich, Germany; E-Mail; Center for International Health, Ludwig Maximilian University of Munich (LMU), Munich, Germany; Division of Infectious Diseases and Tropical Medicine, LMU University Hospital Munich, Germany, Leopoldstrasse 5, D-80802 Munich, Germany; E-Mail; German Center for Infection Research (DZIF), partner site Munich, Germany; Helmholtz Zentrum München, Ingolstädter Landstraße 1, 85764, Neuherberg, Germany; Departments of Mathematics and Life Sciences, Technical University of Munich, Germany; Max von Pettenkofer Institute, Faculty of Medicine, LMU of Munich, Germany

**Keywords:** COVID-19, pandemics, Coronavirus Infections/epidemiology, panel study, Enzyme-Linked Immunosorbent Assay, Models, Economic, Stress, Psychological, socio-economic factors, spatial analysis, geographic information systems

## Abstract

**Background:** The SARS-CoV-2 pandemic is leading to the global introduction of public health interventions to prevent the spread of the virus and avoid the overload of health care systems, especially for the most severely affected patients. Scientific studies to date have focused primarily on describing the clinical course of patients, identifying treatment options and developing vaccines. In Germany, as in many other regions, current tests for SARS-CoV2 are not being conducted on a representative basis and in a longitudinal design. Furthermore, knowledge about the immune status of the population is lacking. Yet these data are needed to understand the dynamics of the pandemic and to thus appropriately design and evaluate interventions. For this purpose, we recently started a prospective population-based cohort in Munich, Germany, with the aim to better understand the state and dynamics of the pandemic.

**Methods:** In 100, randomly selected constituencies out of 755, 3,000 Munich households are identified via random route and offered enrollment into the study. All household members are asked to complete a baseline questionnaire and subjects ≥14 years of age are asked to provide a venous blood sample of ≤3 ml for the determination of SARS-CoV-2 IgG/IgA status. The residual plasma and the blood pellet are preserved for later genetic and molecular biological investigations. For twelve months, each household member is asked to keep a diary of daily symptoms, whereabouts and contacts via WebApp. If symptoms suggestive for COVID-19 are reported, family members, including children <14 years, are offered a pharyngeal swab taken at the Division of Infectious Diseases and Tropical Medicine, LMU University Hospital Munich, for molecular testing for SARS-CoV-2. In case of severe symptoms, participants will be transferred to a Munich hospital. For one year, the study teams re-visits the households for blood sampling every six weeks.

**Discussion:** With the planned study we will establish a reliable epidemiological tool to improve the understanding of the spread of SARS-CoV-2 and to better assess the effectiveness of public health measures as well as their socio-economic effects. This will support policy makers in managing the epidemic based on scientific evidence.

## Background

Since the first description of the novel coronavirus disease (COVID-19) in December 2019 in Wuhan, China, the disease has spread worldwide and was classified as a global emergency by the WHO in early 2020 (1). In Germany, the first confirmed case of COVID-19 was registered on January 6 2020 at the Division of Infectious Diseases and Tropical Medicine, LMU University Hospital Munich (2, 3). The transmission chains were interrupted by contact tracing and isolation of the affected persons. However, due to the return of German tourists from holidays in the high-risk areas of northern Italy in connection with a carnival celebration in the district of Heinsberg (60 kilometres west of Cologne), the virus spread to 13 of 16 federal states within one month (4). The exponential increase in newly confirmed cases in Germany reached a total of 155.193 positively tested cases on April 27 2020 (187 per 100,000 inhabitants) (4).

Simulations and experiences of other countries suggest that healthcare systems would be overburdened and eventually collapse due to a pronounced increase of patients needing intensive care support if no interventions were taken (5–13). In the absence of vaccinations and specific treatment options, public health interventions were initiated in Germany, similarly to numerous other countries comparably affected. The measures include isolation of confirmed patients, quarantine of their contacts, use of personal protective equipment, social distancing (including school closures), and closure of borders (6, 14). Prediction models and experiences from countries like South Korea suggest that combination of these measures could be effective in combatting the disease (13, 15–18). However, past evidence from other epidemics was not that convincing with respect to controlling virus spread by social distancing (19). It remains unclear how comparable previous viral disease outbreaks are to SARS-CoV-2 (20). While potentially saving lives and protecting healthcare systems from breakdown, one has to bear in mind that measures of social distancing can have a devastating impact on national and global economies, healthcare systems, incomes of individuals and families (especially those in precarious employment conditions), education (which particularly affects disadvantaged groups) and on health and the psychosocial well-being of populations (20–22). Devastating effects seen in high-income societies will likely be much worse in low and middle income countries (23).

Results of simulation studies existing so far differ considerably. This is partly due to the unknown number of asymptomatic or minimally symptomatic SARS-CoV-2 carriers, and thus the number of undetected cases (4, 7, 13, 17, 24, 25). In addition, the number of confirmed cases depends on access to healthcare, laboratory availability and on the applied criteria for who should be tested. Therefore, the basic and the effective reproduction number can only be very roughly estimated and the hospitalization and mortality rates remain to be confirmed. Community cohorts can help to assess the overall spread of infection in the community and thus provide more reliable estimates of the basic and the effective reproduction number. This will help to evaluate the burden on the healthcare system as well as the effectiveness of public health interventions (26).

## Methods

### Aim of KoCo19 (Prospective Covid-19 Cohort Munich)

With the community-based household study presented in this paper, we therefore aim to study the sero-prevalence and -incidence of SARS-CoV-2 antibodies in a representative household sample of the Munich population. With this approach we will provide a constantly updated epidemiological instrument that represents the number of infections that have occurred in the city. The study may also serve as a pilot for studies in other areas of Germany and other countries.

The following study questions will be addressed:

1. Baseline visit

- What is the SARS-CoV-2 antibody prevalence in the Munich general population?
- How many of the initially seropositive individuals in the baseline-study were previously tested by pharyngeal swab and nucleic acid amplification (PCR) (positively or negatively) and/or had symptoms suggestive for COVID-19 (yes or no)?
- What is the distribution of symptom severity in each of the groups described above?
- How high is the risk of infection for other household members if one person is infected and can household risk factors be identified?
- How high is the risk of infection for other inhabitants of the same apartment building if one person is infected?
- What are the risk factors for SARS-CoV-2 infections?
2. Follow-up visits

- Is there a change in antibody titers of those who initially tested positive, which is potentially necessary to discriminate from cross-reactivity with other corona viruses?
- How long are SARS-CoV-2 specific antibodies detectable after infections of varying severity?
- How does the spread of the disease develop and what influence do public health measures have on the incidence?
- What is the impact of individual behaviour on the incidence of infection?
- Which risk factors are associated with SARS-CoV-2 incidence?
- What is the socio-economic impact of the pandemic and the measures to combat it, especially on the employment situation and psychosocial endpoints?

### Setting of KoCo19

Munich, the capital of the Free State of Bavaria, is located in the southeast of Germany. Approximately 1.5 million people live here, 9% of these are 75 years and older (27). The population density is 50 inhabitants per ha (27). There are 70 hospital beds (including 5 intensive care beds) and 13 doctors for every 10,000 inhabitants. (27). After the first 100 cases of SARS-CoV-2 were reported in Munich by March 12 2020, the Bavarian schools and universities were closed on March 16 2020, initially until May 11 2020. Since the same date, all shops that do not serve the basic needs of the population were closed. Starting on March 21 2020, when 1,288 infected individuals were reported in Munich, curfews were implemented (28). According to these curfews, people are essentially only allowed to leave the house to go to work, to the doctor, to buy food, for outdoor exercise (jogging, walking) or to help others who are depending on support. A minimum distance of 1.5 m to other persons must be maintained.

### Design of KoCo19

The study design of KoCo19 is a community-based prospective cohort study in randomly selected Munich households. All members of the selected households who are eligible and agree to participate (see “Study population”) are invited to the following parts of the study:

1. **Baseline study (1^st^ household visit)**: During the baseline study, personal identifying information is collected and stored in a database separately from the remaining questionnaire information. A blood sample is taken from which sero-prevalences of SARS-CoV-2 IgG and IgA antibodies are determined. After the household visit, participants are asked to answer an:

a. **Online household questionnaire** and an
b. **Online personal questionnaire**.
2. **Daily diary**: Using a web-based app, participants are asked to complete a daily diary on symptoms suggestive of COVID-19 infection, whereabouts and social contacts. Additional questions might be included throughout the follow-up period. If symptoms of COVID-19 occur, a pharyngeal swab for PCR testing of SARS-CoV-2 is offered at our division.
3. **Follow-up household visits**: Households are re-visited every three to six weeks for a new blood sample in order to estimate the sero-incidence of SARS-CoV-2 infection. This frequency can be adapted to the current necessities of estimated prediction models. The follow-ups are currently planned for up to 12 months.

The study will be terminated if more efficient methods to assess the course of the epidemic are developed or if this no longer appears relevant.

### KoCo19 Study population

For KoCo19, a representative sample of Munich households (target population) is selected by random walk door-to-door methodology (29). For this purpose, 100 of the 755 Munich constituencies were randomly selected using R (The R project). In each of these constituencies, the geographic center is selected as the starting point of the random route using QGIS (Figure 1). From the address closest to this starting point, 30 households per constituency will be included in the study according to a fixed algorithm. In the case of apartment buildings, one household per floor is selected to investigate possible transmission within the house.

**Figure 1:**
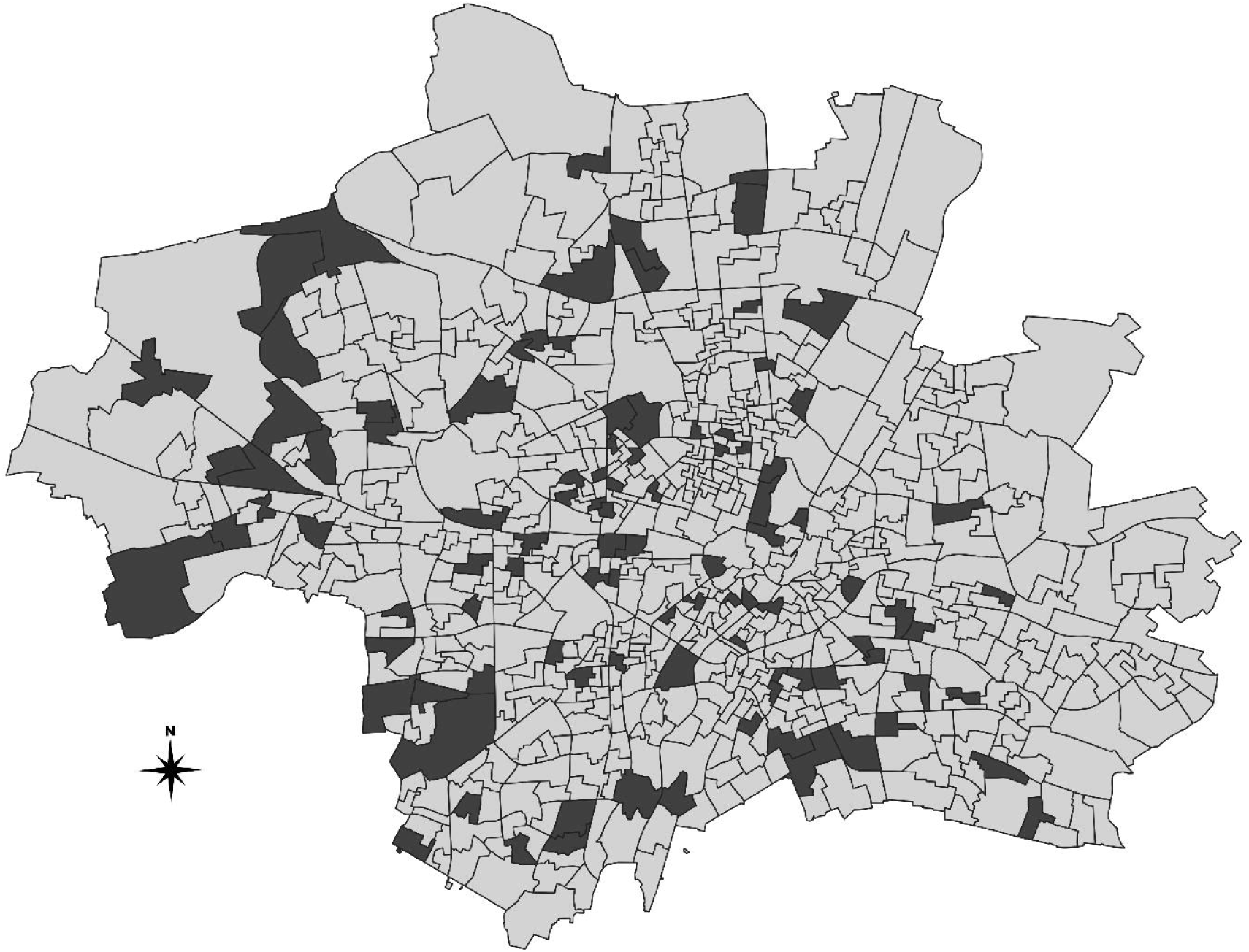
Distribution of the selected 100 out of 755 Munich constituencies. Each of the 20 teams of field workers is responsible for recruiting 30 households in five constituencies following a random route algorithm (© Landeshauptstadt München – Kommunalreferat – GeodatenService - 2020).

All household members ≥14 years are invited to participate in KoCo19 by donating a maximum of 3 ml of blood and to be available for further blood tests every three to six weeks. Participants are informed about their SARS-CoV-2 antibody status. Additionally, all household members are asked to complete a daily questionnaire on their state of health, whereabouts and social contacts using an internet or a smartphone app (WebApp). Persons who do not have a mobile phone or cannot operate an App will be interviewed by phone. At least one household member needs to agree to donate blood, while other members can solely participate in the questionnaires.

Inclusion criteria are:

- At the time of inclusion into the study (1^st^ household visit), at least one of the household members must be ≥18 years and competent to provide written informed consent.
- Sufficient command of German to understand the participant information materials for the study and to answer the questionnaires (Note: Due to the urgency of the study, there is no possibility to develop respondent information and questionnaires multilingually or to recruit multilingual study teams).

Households where the residents are not present at the time of the study team’s visit and do not call back to arrange a baseline visit, do not give informed consent or who do not meet the inclusion criteria will be

- replaced by the next house on the route for single/two-family houses
- replaced by the next apartment on the same floor in the case of apartment buildings.

Non-response is noted and taken into account in the calculation of the response index. Where feasible, basic information (age, sex, type of building) and reason of non-participation are collected for non-responders in order to assess representativeness of the study population. In addition, participants’ socioeconomic status, migrant status, sex, and percentage of households with children and single households will be compared to the official statistics of the selected constituencies and of all Munich constituencies (Statistical Office Munich).

### Field work

In order to pre-inform the population about the study and thus, increase response, the study is announced in the media and on a webpage (www.koco19.de) including an information video (https://youtu.be/O_Qznp8FEA8). In addition, field workers visit the selected households before the baseline study to introduce the study, hand out a short leaflet and complete study information. In case of absence at the time of the information visit, the teams leave information material including a telephone number in order to schedule the baseline visit. For the information visit, teams are accompanied by a police officer; this is considered helpful to enhance trust in the study in times were reportedly fraudsters are taking advantage of the exceptional situation.

Overall, at least 50 field workers working in 25 teams of two are involved in the study. Each team is responsible for 150 households in five constituencies (figure 1). One field worker is a medical student with prior, extensive training in infectious disease control, including blood sampling and pharyngeal swabs in case participants have symptoms suggestive for COVID-19 within 14-days prior to this visit. The second field worker is responsible for informed consent and interviews. Teams of field workers are carefully trained in study procedures, data confidentiality, and infection protection and undergo a proficiency test before initiating field work. During the first field visits, they are accompanied by a senior medical doctor of the Division of Infectious Diseases and Tropical Medicine, LMU University Hospital Munich, until the physician approves correct handling of all steps of the field work. To further ensure the quality of field work randomly selected households are called and asked about the last study visit and potential problems. In addition, teams will be repeatedly monitored by a senior medical doctor throughout the study. To avoid infection risks through public transport, all teams use rental cars during the field work, to carry all the necessary material including personal protective and hygiene equipment.

### Study instruments: Questionnaires

Wherever possible, questions were taken from pre-existing validated questionnaire instruments (30–33). As it will be crucial to minimize attrition over time, we minimized the number of questions without losing important information.

#### - Household questionnaire

The household questionnaire includes questions on the living situation (type of housing, number of bedrooms, apartment size), number of inhabitants (including date of birth and sex), highest level of education, work situation, household income, second hand smoke exposure, work of household members in potentially high risk jobs for SARS-CoV-2 infections, past pharyngeal swab testing for SARS-CoV-2 in household members including test results.

#### - Individual baseline questionnaire

At baseline, all participating household members are asked about date of birth, sex, level of education, employment situation, smoking history, general health, pregnancy, recent influenza vaccination, pre-existing medical conditions, symptoms suggestive for COVID-19 in the 14 days prior to the study, past PCR testing of nasopharyngeal samples for SARS-CoV-2 including test result, use of respiratory masks, and work in a potentially high risk job for SARS-CoV-2 infection.

#### - Diary

The daily diary includes items about symptoms suggestive for COVID-19, social contacts, whereabouts and use of public transport in the past 24 hours. Further questions on e.g., the psychosocial and economic situation, such as perceived health status, behavioral aspects, or employment and income will be added over the time of the study, and collected e.g. once a week.

### Laboratory analyses

Samples will be analyzed and stored at the Division of Infectious Diseases and Tropical Medicine, LMU University Hospital Munich.

First, blood is sampled in 2.7 ml EDTA containers and thoroughly mixed. Samples are individually barcoded and packed to be transported to the laboratory in 12 V car battery powered coolers. There, the samples are centrifuged to separate the cell pellet from the remaining plasma. Cell pellets are frozen at -80°C for further analysis, while the plasma is used for ELISA analysis using a semi-automated robotic system (Euroanalyzer I, Euroimmune, Lübeck, Germany). Serology is performed primarily using the Anti-SARS-Cov-2-ELISA IgG and IgA (Euroimmune, Lübeck, Germany). The ELISA system has a combined sensitivity of between 66.7% (<10 days after onset of symptoms) and 100% (> 10 days after onset of symptoms) according to the manufacturer. Specificity is rated as 98.5%, tested in larger cohorts of blood donors. The remaining plasma is stored for further analysis or confirmatory testing e.g. with virus neutralization as appropriate.

Pharyngeal swabs are taken using eSwab systems. The samples are stored at 4°C and immediately transported to the laboratory. There, RNA extraction is performed. Extracted RNA is divided to allow for cryo-conservation at -80°C as well as for diagnostic RT-PCR for SARS-CoV-2. The reserve sample will be used for virus sequence analysis to perform cluster and outbreak analysis and to study within family transmission.

## Statistical analysis

### Sample size calculation

As an initial number of participants, 3,000 households with approx. 1.5 participants each were calculated. Assuming that each person is included in the study with the same probability, repeated drawing of 4,500 from 1.5 million Munich residents will yield the 95% confidence intervals listed in Table 1 for the subsequently assumed prevalence of the total of reported and unreported infections in the baseline survey. The prevalence of confirmed cases was 0.3% on April 9 2020 (34). This shows that the sample size is sufficient for an adequately precise estimate of the actual sero-prevalence in the baseline survey.

**Table 1:** 95% Confidence Intervals of SARS-CoV-2 prevalence in the study population by assumed true prevalence in the Munich population (The R Project)

| Assumed true prevalence of<br>SARS-CoV-2 seropositive<br>residents of Munich<br>(1.5 million inhabitants) | 95% Confidence Interval of<br>SARS-CoV-2 seropositive participants<br>in KoCo19<br>(N=4,500) |
| --- | --- |
| 0.5% | 0.31% - 0.71% |
| 1% | 0.71% - 1.29% |
| 5% | 4.38% - 5.64% |
| 10% | 9.13% - 10.89% |
| 20% | 18.84% - 21.18% |
| 30% | 28.67% - 31.31% |
| 50% | 48.56% - 51.47% |

### Data management

Data will be stored and handled at the Division of Infectious Diseases and Tropical Medicine, LMU University Hospital Munich. The pseudonymized databases will be combined using a unique participant ID. Using the combined raw data, a scripted routine analysis is used to produce a daily update of descriptive and bivariate prevalence and incidence data to generate a study “dashboard” (The R project). Main outcome variables are the prevalence and incidence of SARS-CoV-2 antibodies as well as SARS-CoV-2 symptoms in the study population and the dynamics thereof. Main exposure variables are socio-economic factors, social contacts, city district as well as the living situation. In addition, changes in the non-pharmaceutical public health interventions will be used as predictor of SARS-CoV-2 incidence.

### Descriptive analyses

Initial descriptive data analyses will be weighted for cluster sampling and include the following parameters:

- Description of basic data for responder and non-responder households
- Socio-demographic data and known risk factors for SARS-CoV-2 infection
- Baseline prevalence of SARS-CoV-2 sero-positivity in the Munich general population stratified for a) symptomatic and asymptomatic cases and b) cases previously tested via PCR
- The temporal course of SARS-CoV-2 sero-positivity in the Munich general population (point prevalence and incidence) stratified for symptomatic and asymptomatic subjects and reported cases
- The daily prevalence and incidence of possible COVID-19 symptoms in the study population

### Bi-variable and multi-variable analyses

Subsequent bi- and multivariable analyses, taking cluster sampling into account, will include the following aspects:

○ Identification of risk factors for asymptomatic, mildly symptomatic and severely symptomatic SARS-CoV-2 infections (age, sex, socioeconomic status, occupation, social contacts, district). Prevalence ratios are calculated for this purpose.
○ The temporal relationship of public health interventions (school closures, etc.) with the incidence of SARS-CoV-2 symptoms and changes in sero-prevalence are analysed, e.g. by using mixed effect models with a time varying covariate indicating the different intervention variables at different time points.
○ The effect of discontinuation of public health interventions (school closures, etc.) are analysed longitudinally, e.g. by mixed effect models.
○ The interaction of sociodemographic, economic and psychological variables with the epidemic and with interventions to contain it will be analysed.
○ An algorithm for the most reliable prediction of SARS-CoV-2 PCR positivity will be developed.
○ Geo-spatial modelling of exposures and outcome will be performed.

In addition, the data is used to gain knowledge about spread dynamics and to predict the further development of the epidemic under different scenarios. Models to be used for this purpose are developed during the course of the study.

## Discussion

The ongoing SARS-CoV-19 pandemic has changed life globally to an extent unseen before. Due to the lack of vaccinations and pharmacological treatment options, it was predicted that not taking public health action will result in an overload of healthcare systems in most countries and a mortality of millions in the global population (25). The public health interventions that have recently been implemented by most countries have a huge impact on the economy and most likely, also on health and well-being of the global population. However, in order to reliably estimate the total number of previously infected individuals (with and without symptoms), who are – hopefully – resistant to infection for an extended period of time, population-based representative household studies are helpful to understand the dynamics of the disease.

The study presented here will provide a first estimate of the prevalence and incidence of sero-positivity in the Munich population. Although not generalizable on a global scale, this will give first insights about the proportion of asymptomatic and mildly symptomatic carriers of SARS-CoV-2 in comparison to the number of those tested. It will also help to identify risk factors for infection, course of disease and effectiveness and efficiency of the public health measures.

Our study has limitations. In the last years, willingness to participate in population-based studies went down considerably (35, 36). Low response might affect representativeness of the study population, which in turn might have an effect on the generalizability of the prevalence of positive antibody results to the Munich source population. However, it is unlikely that participation will depend on sero-positivity of antibody results as antibody status is unknown prior to inclusion in the study. In addition, the research topic is of uppermost interest for many citizens in the current situation, therefore response is expected to be higher in KoCo19 than in other studies. During the first recruitment days, an overall response of close to 50% was reached providing some evidence for this hypothesis. Response will be increased by revisiting households which did not open the door at the first visit. For the associations under study, representativeness is of less concern (37). However, we might not be able to reach high response especially in specific groups of the target population, e.g. subjects with migration background as they are generally harder to reach in epidemiologic studies (38) and because time constraints do not permit to provide study documents in other languages than German at the initiation of the study. Likewise, the spread of SARS-CoV-2 varies locally and depends on several factors, such as the time course of the infection in the respective region, the population density and age distribution of the population, the available capacities and the countermeasures taken. Therefore, prevalence and incidence results obtained in this study locally might not be easily generalizable to other cities, regions or countries. Losses-to-follow up will likely occur especially when public health interventions are relaxed and the public attention focuses less on the pandemic. However, as long as these missing data can be assumed to be missing at random multiple imputation can account for attrition (39).

With respect to reporting bias, one may assume that participants are more likely to over-report symptoms which will result in an overestimation of the symptom prevalence. In order to minimize other forms of reporting bias we use as many items from validated questionnaire instruments as possible. In addition, all items were carefully checked by several members of the KoCo-19 team for their face validity. Questionnaires, especially the daily diary, were kept as short as possible. Therefore, the study might not be able to answer specific questions in favour of more valid answers and low attrition rate over time. For such aspects, case-control studies, potentially nested in the current study, could be done. For ethical reasons, participants will receive their SARS-CoV-2 antibody status after each blood sampling. This might influence their subsequent behaviour, especially when antibody status is positive.

To minimize social desirability bias, questionnaires are web-based and to be completed by the participants themselves. However, as not all participants might have web-access and especially older participants might not have the necessary internet competencies, a telephone interview is also offered. Type of response (online or interview) is being recorded so that systematic differences in response can be accounted for. We are not able to include children under the age of 14 years from the beginning mainly due to ethical concerns regarding the venous blood sampling by a medical student. Over the course of the study development of other test methods might allow the inclusion of this important part of the population.

Random route recruitment is a feasible way of recruitment where population lists do not exist or are hard to obtain. It has been applied by the WHO in various vaccination studies since the 1980s, was modified over time and it is also used in large scale community surveys such as the European Working Condition Survey (29, 40). The alternative approach, sampling via the Munich population registry, would have taken more time due to the formal requirements and thus would have slowed down the start of the study. As we apply a cluster sampling approach (100 out of 755 constituencies) and include more than one person per household and more than one subject per apartment building, clustering has to be taken into account in the statistical analyses. Taking constituencies made inclusion of 3,000 households and the follow-up visits within six weeks feasible. Inclusion of more than one household member and one household per floor will help us to better understand the spread of SARS-CoV-2 within households and apartment buildings. In addition, inclusion of more than one household per apartment building takes into account that larger number of inhabitants live in apartment buildings than in one- or two-family houses, which would otherwise have been overrepresented.

The sample size calculation reported here did not take clustering into account. This was due to the fact that the prevalence of SARS-CoV-2 within clusters (household, apartment building, constituency) is so far unknown. Taking a more conservative approach of considering only 3,000 participants (= number of households) instead of 4,500 participants, the 95% Confidence Interval for a given prevalence of 0.5% (50%) would increase from 0.3–0.7% (49–51%) to 0.3–0.8% (48–52%). Thus, changes are minimal and we therefore conclude that our prevalence estimates will be precise.

Currently there is only a limited number of reliable serologic tests available. By using the – to our knowledge – best currently available serology test system with IVD certification in the study, the number of valid results will be maximized as compared to other less mature testing systems. Still, with very low sero-prevalence, the false positive rates in the population might be in the range of the test-background. All currently performed studies face this limitation, however by repeated visits, the seroconversions will be confirmed, thus offering much better data than mere sero-prevalence data. Besides, it could not yet be established that sero-positivity in ELISA corresponds reliably with immunity. Thus further testing such as virus neutralization is performed for questionable cases.

As serology only allows the detection of infection in retrospect, the pharyngeal swab is essential to pick up acute infections. This also allows to detect subjects with symptomatic disease who possibly never develop positive serology; although it is currently believed that most patients will develop positive serology within 10 days after onset of symptoms. Besides, the swab can be used to extract viral RNA and use it for sequencing, offering further information about transmission dynamics with households, quarters or even worldwide.

In a pandemic situation, it would neither be ethical nor feasible to use medical doctors for epidemiological field work as they are needed for clinical service. Therefore, experienced medical students together with students of other subjects perform the field work of this study. The medical students involved are not removed from other important tasks during the pandemic. Ethical considerations are also relevant for the use of personal protective equipment in this study. Currently, personal protective equipment is available at Munich hospitals. Its’ availability is being re-evaluated regularly over the duration of the field work and the demand for personal protective equipment in clinical services will always have priority.

### Conclusion

KoCo19 is a unique possibility to obtain more reliable estimates of the spread of SARS-CoV-2 in the general population and to better understand the dynamics of COVID-19. Although a single epidemiologic cohort study in one city will not be able to answer all questions related to SARS-CoV-2; it will provide an important epidemiological basis for our understanding of the epidemic, and might serve as a blueprint for similar studies.

## Declarations

### Ethics approval and consent to participate

The study protocol was approved by the Institutional Review Board at the Ludwig-Maximilians-University in Munich, Germany (opinion date 31 March 2020, number 20–275). Written informed consent is sought from all participants and, in case of minors, by their legal guardian(s). The pseudonymization of the data enables us to provide the participants with individualized results. Personal identifying information is stored in a password-protected database containing only contact details, contact status and contact IDs. The data for each questionnaire response and the other samples are stored in a separate password-protected database containing only ID numbers. It is therefore not possible to draw any conclusions about the identity of the participants and the information they have provided. Compliance with data protection regulations was approved by the official data protection officer of the University Hospital of the LMU Munich, Gerhard Meyer. The data are stored for as long as they are needed to achieve the study objectives. Ten years after the start of the study (and every three years thereafter), the study team will evaluate whether the study objectives for the project have been achieved and, if so, will request the deletion of all personal data.

## Data Availability

Data of this is not yet available as this is a study protocol.

## Consent for publication

Not applicable

## Availability of data and materials

Not applicable

## Authors contribution

MH is the principal investigator of the study and obtained the necessary funds. Together with KR and ES, MH designed the study and wrote the study protocol. MP, JMGN, IK, LO, FR and VT conceptualized the field work and made significant contributions to the study protocol. MD is responsible for the conception of all IT related study components. FF is coordinating the data management. FT conceptualizes the modelling of the spread and dynamics of the disease. AW is responsible for the laboratory methods of KoCo19. All authors approved the submitted version of the paper and have agreed to be personally accountable for the author’s own contributions and to ensure that questions related to the accuracy or integrity of any part of the work, even ones in which the author was not personally involved, are appropriately investigated, resolved and the resolution documented in the literature

## Acknowledgements

We thank the KoCo19 advisory board Prof. Dr. Stefan Endres, Prof. Dr. Matthias Tschöp, Prof. Dr. Andreas Zapf, Prof. Dr. Manfred Wildner, and Stefanie Jacobs. We thank Accenture for the pro-bono development of the KoCo19-Survey-Webapp.

We are grateful to our students at the KoCo19 coordination office: Jan Marius Bruger, Simon Michael Winter Jared Anderson; our teams of field workers: Paula Matcau, Norah Kreider, Emma Dech, Laura Charlotte Dech, Tim-Oliver Haselwarter, Julian Ullrich, Isabel Klugherz, Kristina Gillig, Kerstin Puchinger, Michael Höfinger, Tobias Würfel, Julius Raschka, Stefan Hillmann, Alexander Maczka, Konstantin Pusl, Isabel Brand, Philine Falk, Alisa Markgraf, Stefanie Fischer, Clemens Lang, Magdalena Lang, Silvan Lange, Claire-Sophie Pleimelding, Leonie Pattard, Tim Hofberger, Sonja Magdalena Gauder, Marius Gasser, Vitus Maximilian Grauvogl, Marco Scholz, Hannah Maria Müller, Jonathan Leon Frese, Valeria Baldassarre, Charlotte Kiani, Anna Ngoc-Nhu Do, Sophie Schultz, Ekaterina Lapteva, Tim Haselwarter, Maximilian Baumann, Elias Golschan, Vitus Grauvogel, Vera Britz, Eva Thumser, Paul Schandelmaier, Patrick Wustrow, Lea Zuche, Janna Hoefflin, Jeni Tang, Julia Wolff, Rebecca Böhnlein, Flora Deak, Lara Schneider, Jakob Reich, Leonard Gilberg, Matthias Hermann, Thomas Zimmermann, Friedrich Caroli, Niklas Thur, Felix Lindner and our telephone interviewers: Nicole Schäfer, Julia Waibel, Pia Wullinger.

## # KoCo19 Collaborators are

### LMU University Hospital Munich

*Division of Infectious Diseases and Tropical Medicine*: Abhishek Bakuli, Judith Eckstein, Günter Froeschl, Otto Geisenberger, Christof Geldmacher, Arlett Heiber, Larissa Hoffmann, Kristina Huber, Dafni Metaxa, Camilla Rothe, Mirjam Schunk, Claudia Wallrauch, Thorbjörn Zimmer

Institute for Emergency Medicine and Medical Management: Stephan Prückner

### Helmholtz Zentrum München

*Institute of Computational Biology*: Christiane Fuchs (main affiliation: Bielefeld University, Faculty of Business Administration and Economics), Jan Hasenauer (main affiliation: University of Bonn, Interdisciplinary Research Unit for Mathematics and Life Sciences), Mercé Gari

*Institute for Radiation Medicine, Integrative Modelling*: Noemi Castelletti

*Institute of Translational Genomics*: Eleftheria Zeggini

*Institute of Health Economics and Healthcare management*: Michael Laxy, Reiner Leidl (also affiliated at: Munich Center of Health Sciences, Ludwig Maximilian University of Munich), Lars Schwettmann

